# Evaluation of a Risk-Stratified National Breast Screening Programme in the United Kingdom: An updated cost-effectiveness analysis

**DOI:** 10.1101/2024.04.16.24305897

**Authors:** Stuart J Wright, Gabriel Rogers, Ewan Gray, Anna Donten, Lorna McWilliams, David P French, D Gareth Evans, Katherine Payne

## Abstract

**Study objective:** To update a published early economic evaluation of exemplar risk-stratified national breast screening programmes (stratified-NBSP).

**Method:** An existing validated decision-analytic model, using discrete event simulation (the ‘Gray-model’), was used to structure the pathways for 3 stratified-NBSP (risk-1; risk-2; risk-3) compared with the current NBSP in the United Kingdom (UK-NBSP), biannual screening, and no screening. The updated model is called MANC-RISK-SCREEN and assumes a life-time horizon, the UK health service perspective to identify costs (using £; 2022) and measures health consequences using life-years and Quality Adjusted Life Years (QALYs). The original data sources used for the Gray-model were assessed for current relevance and updated where feasible. Updated data sources included: cancer and all-cause mortality; breast cancer incidence; breast cancer risk data; tumour staging; recall rate; mammographic sensitivity by breast density group; costs; and utilities. Model parameter uncertainty was assessed using Probabilistic Sensitivity Analysis (PSA) and one-way sensitivity analysis.

**Results:** The base case analysis, supported by PSA, suggested that there was always a risk-stratified approach to breast cancer screening that was superior to universal screening. In the base case analysis, a strategy of dividing women into three equal groups based on risk was the most cost-effective. In the PSA, a strategy based on that used in the BC-PREDICT study was the most cost-effective. There was uncertainty in whether the addition of reduced screening for women at lower risk was cost-effective.

**Conclusion:** The results of this study suggest that risk-stratified approaches to breast cancer screening are more cost-effective than both 3-yearly and 2-yearly universal screening.

**Highlights:**

- A published early decision-analytic model-based cost-effectiveness analysis, using discrete event simulation (the ‘Gray model’), produced indicative results suggesting all included exemplars of a stratified national breast screening programme (stratified-NBSP) were cost-effective compared with no screening but a fully incremental analysis indicated only risk-based stratified-NBSP were cost-effective.
- This study uses a subsequently validated version of the Gray-model to produce a cost-effectiveness analysis with an updated model called MANC-RISK-SCREEN using revised descriptions of the relevant stratified-NBSP and new values for cancer and all-cause mortality; breast cancer incidence; breast cancer risk data; tumour staging; recall rate; mammographic sensitivity by breast density group; costs; and utilities.
- This analysis builds on the indicative estimates of the healthcare costs and health consequences of stratified-NBSP and suggests, with the current level of evidence, they are a cost-effective use of the NHS budget in the United Kingdom but uncertainty remains in the value of reducing screening for those at lower risk.

## Introduction

There is emerging evidence to indicate the potential value of using stratified approaches in national breast screening programmes (hereafter ‘risk-stratified-NBSP’) ^1,2^. A risk-stratified-NBSP would use information from a risk-prediction tool, such as the Tyrer-Cuzick questionnaire, to allocate women to a risk category (e.g. population risk; low risk; high risk; very high risk) using predefined thresholds ^3^. The risk category is then combined with a pathway of care to produce a screening programme using mammography (and/or other diagnostic tools) at defined screening intervals to identify tumours ^4^. The primary motivation for implementing a stratified-NBSP is to improve the ratio of benefits, harms and healthcare costs resulting from screening by adjusting the time between screening intervals to match the estimated risk of developing a tumour causing breast cancer ^5^. Women that are allocated to a high-risk category will be invited to more regular screening and cancer-preventing drugs when compared with women at population risk or low risk of breast cancer.

National bodies across Europe are starting to collate evidence to support whether to replace the existing, ‘one size fits all’ approach to breast screening programmes ^6^. A number of studies have demonstrated the potential feasibility of risk-stratified breast cancer screening ^7–10^. In the UK, the current NBSP involves 3 yearly screening from the age of 50 to 70. Any subsequent modifications to a NBSP will have an opportunity cost and in the UK, the National Screening Committee is clear about the need for evidence on the benefits, harms and cost-effectiveness of any new programme ^11^.

The estimated cost of a risk-stratified NBSP in the UK has been calculated using micro-costing methods to be £6.64 per woman, generating a UK-population cost of £1,460,800 per year (at 2021 prices) ^12^. To date, three published economic analyses of exemplar risk-stratified-NBSP are relevant to the UK setting. Pashayan and colleagues focused exclusively on the effect of reducing the time between screening for women not at high risk of breast cancer, with varying thresholds used to define high risk ^2^. A recent study by Hill et al found that risk-based breast cancer screening strategies based on increasing screening for those at high risk and reducing screening for those at lower risk were likely to be cost-effective in the UK ^13^. However, a limitation of the Hill et al model was that it was not underpinned by tumour growth model and tumour stage was assigned independently of tumour size.

In 2017, Gray and colleagues published an early decision-analytic model-based cost-effectiveness analysis focused on understanding the indicative healthcare costs and health consequences of exemplar stratified approaches for use in a NBSP ^1^. Their model (hereafter ‘the Gray-model’), which includes a model of breast tumour growth, has subsequently undergone validation, including complete technical verification ^14,15^. This study aimed to update the early analysis published by Gray and colleagues using the validated form of the decision-analytic model to identify and quantify the healthcare costs and health consequences of three exemplar risk-stratified-NBSP compared with the current NBSP (3-yearly screening), an NBSP with biennial screening (as is common in most European programmes), and no screening. This updated model-based cost-effectiveness analysis was motivated by the recommendations from Sculpher and colleagues advocating the role of iterative economic evaluation to inform budget allocation decisions ^16^.

## Methods

An existing early model-based cost-effectiveness analysis, using the Gray-model, was updated to address the key criteria described in Table 1. The Gray-model underwent validation and modest amendments to its structure; for details, see Wright et al ^14^.

**Table 1:**
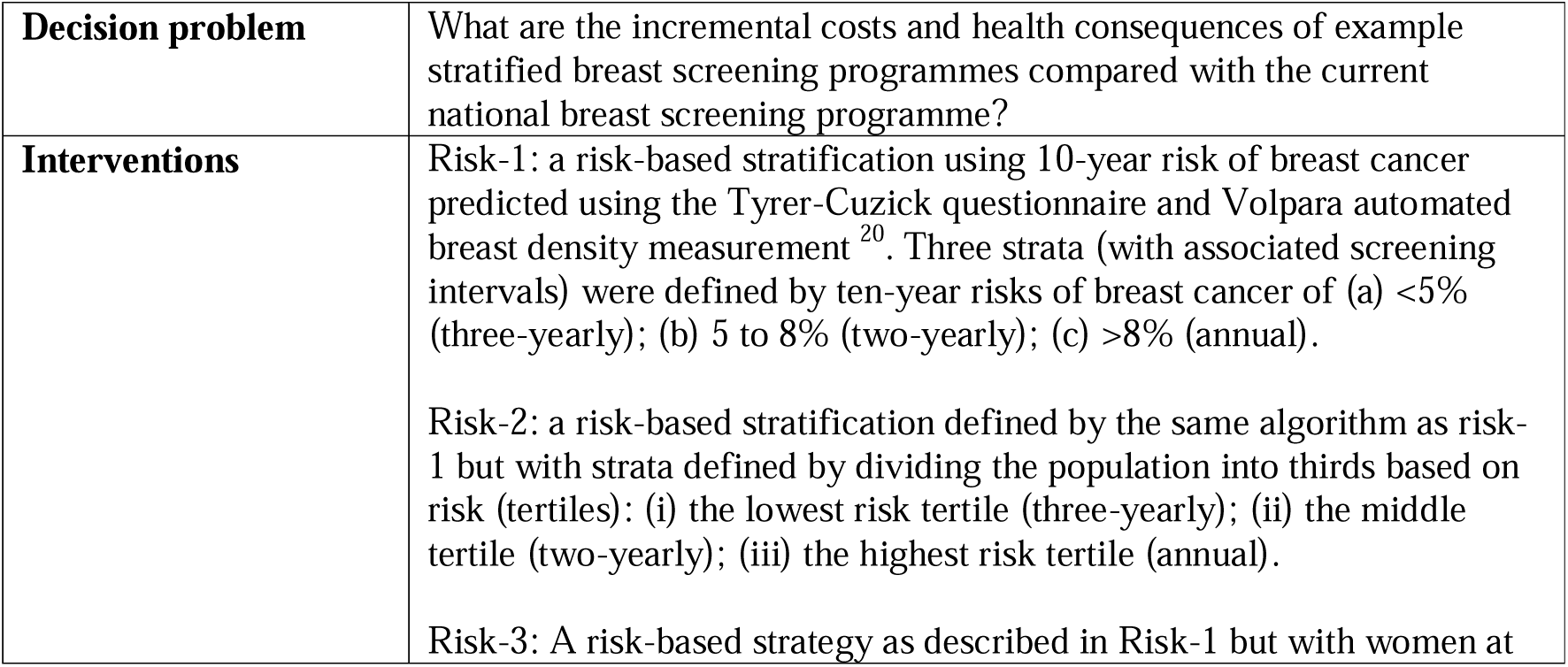
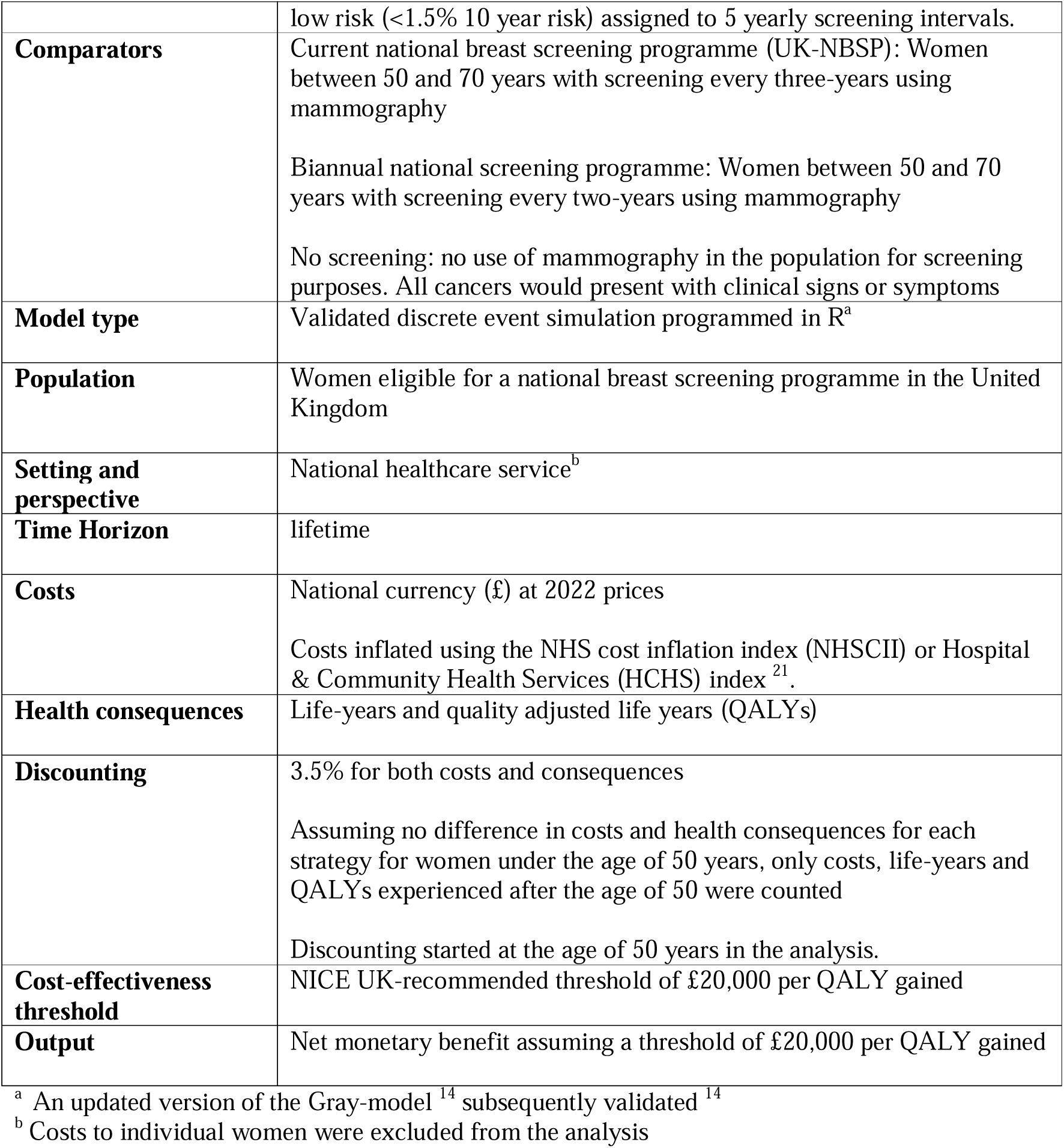
Key design criteria of MANC-RISK-SCREEN.

The Gray-model code and the updated analysis were built in the R statistical package ^17^. The updated Gray-model is called MANC-RISK-SCREEN and is available in a publicly available GitHub repository ^18^. The reporting in this paper follows published criteria ^19^.

### Interventions

The Gray-model assessed four potential approaches to risk that were developed as part of a European Commission funded project (ASSURE: Adapting Breast Cancer Screening Strategy Using Personalised Risk Estimation) ^5^. These are described in Supplementary Appendix 1.

The present analysis modified the relevant interventions included in the Gray-model to more closely align with the suggested approaches, informed by a programme of work looking at the feasibility of rolling out risk-stratified NBSP into the NHS (called PROCAS-2)^22^(p2)^,23^.

The PROCAS-2 programme study sought to evaluate the feasibility of implementation of a risk-prediction strategy at a woman’s first attendance at breast cancer screening. The chosen approach, BC-PREDICT, used an online version of the Tyrer-Cuzick risk questionnaire to capture information on women’s family history, demographic, and lifestyle risk factors. This information was combined with automated breast density measurements captured at the woman’s first mammography appointment using the Volpara TruScan system. Using these two instruments, women’s 10-year and lifetime breast cancer risk were estimated.

Importantly, this feasibility study did not consider strategies with supplemental screening for women with denser breasts as these were found to be unlikely to be cost-effective in the Gray model. The full process of identifying breast cancer risk and providing risk feedback is detailed in French et al 2020 ^20^.

We considered three risk-stratified approaches to breast cancer screening; see Table 1 for details, including the thresholds used to define risk groups. In the first strategy (‘Risk 1’), women estimated to have a high risk of breast cancer are invited to annual screening, those with a moderate risk to biannual screening, and all other women to 3-yearly screening. This strategy aligns most closely with current NICE guidelines for offering addition screening and prevention to women at higher risk of breast cancer ^24^. Strategy Risk-2 divides women into tertiles based on their 10-year risk with those in the highest third attending annual screening, those in the middle third attending biannual screening, and those in the bottom third attending 3 yearly screening. Risk-3 is identical to Risk-1 but with women predicted to have low breast cancer risk invited to less frequent (5-yearly) screening.

### Comparators

Consistent with the Gray-model, we compared risk-stratified approaches with the current UK-NBSP and no screening (definitions shown in Table 1). An additional comparator was added representing an unstratified NBSP that uses biannual screening for all women, which is the approach used in many European countries such as The Netherlands ^25^.

### The relevant population

The relevant population for this decision problem (see Table 1) is women aged 50 years followed until death or age 100 years. Women are simulated from the age of 38 years to allow time for tumours to develop before the screening programme begins. The model code for the Gray-model was re-structured to reduce the total number of simulated women in the population (population size of 10 million in the Gray-model) that was required to achieve a stable rank ordering of the cost-effectiveness of the strategies. The size of the required simulated population was reduced to 2 million women aged 38 years or over by pre-creating a data frame of women who could be simulated through each of the screening strategies rather than drawing new individuals for each strategy. Women who develop cancer before aged 50 years, or who die from other causes before aged 50 years, are removed from the sample. This significantly reduces the underlying variation caused by the simulation model.

### The model structure

The Gray-model simulates individual women moving through the chosen screening strategy using a discrete event simulation structure built around four components: the stratification process; cancer screening; cancer diagnosis and treatment by cancer stage; death. In addition, the model takes an algebraic approach to the natural history of cancer progression following the identification of a tumour. Key design criteria of MANC-RISK-SCREEN are presented in Table 1. For a detailed description of the model structure and diagrams illustrating MANC-RISK-SCREEN, see the documentation folder in the GitHub repository ^18^.

The component parts of the MANC-RISK-SCREEN decision-model are now described in brief, including if, and how, they were modified as part of the validation and update of the Gray-model. During the technical verification process conducted during model validation, some minor errors were also identified in the model code that were rectified ^14^.

#### The stratification process

Consistent with the approach taken in the Gray-model, each simulated woman is assigned a Volpara breast density score, 10-year risk, and lifetime breast cancer risk based on data from a published observational study ^7,23^. The Gray-model used data from the original PROCAS study where risk was predicted for 50,000 women using the Tyrer-Cuzick questionnaire and Volpara breast density measurement. This updated analysis uses data from a second observational study, BC-PREDICT where risk data was calculated for 15,613 women using updated versions of the Tyrer-Cuzick questionnaire and Volpara breast density measurement^20,26^.

#### Cancer screening

Based on her estimated 10-year risk, each simulated woman is assigned to different screening intervals. Changes in the risk groups used in recently published clinical research meant that in the updated model the risk thresholds used to define the different risk-groups have changed^20^. In the Gray model, moderate risk was defined as a 10-year risk between 3.5% and 8%. In MANC-RISK-SCREEN, moderate risk is defined between 5% and 8% in the PROCAS-2 based strategies. This also means that normal risk is defined as less than 5% in the first PROCAS-2 strategy. In the Risk-3 strategy with less frequent screening for women at lower risk, normal risk is now defined as a 10-year risk between 1.5% and 5%.

For each screening event, costs are added reflecting the technology used. The cost of risk-stratification is added at the first screen. A given proportion of women not diagnosed with cancer experience a false-positive result requiring follow-up and potential biopsy, with associated costs added when these occur.

#### Natural History of Cancer

The model represents the natural history of cancer using a mathematical algorithm (see the model diagrams and tumour growth model functional form choice documents in the GitHub repository) ^18^. A random number is drawn and compared with the woman’s lifetime cancer risk to determine if she will experience a cancer in her lifetime. If so, the age of clinical diagnosis is drawn from a distribution of diagnosis ages. A tumour growth rate is drawn and the age of tumour genesis back-calculated using a tumour-growth model ^27^.

At a screening event, the size of the tumour (if present) is determined using the tumour growth model.

The sensitivity of screening is conditional on the size of the tumour and the woman’s breast density. A random number is drawn against this sensitivity to determine if the tumour is detected. The tumour-growth model and sensitivity of screening conditional on tumour size are the same as used in the Gray-model.

#### Diagnosis and treatment

When a tumour is diagnosed through screening or by reaching the size at which it would be clinically diagnosed in the Gray-model, a Nottingham Prognostic Index (NPI) rating is assigned using the distribution of NPI ratings for different tumour sizes. A fixed proportion of cancers are allocated to be ductal carcinoma in situs. MANC-RISK-SCREEN now uses TNM (extent of the tumour (T); extent of spread to the lymph nodes (N); presence of metastasis (M)) stages instead of NPI due to the larger availability of data for this grading system ^28^.

When a woman is diagnosed with breast cancer, a lookup table is used to assign the total cost of treatment given the stage of cancer, age at diagnosis, at number of years the woman will live with breast cancer ^29^. It is assumed that there are no additional treatment costs after the ninth year.

#### Mortality

In the Gray-model, age of death is drawn using an exponential survival function for cancers of the given NPI rating. In MANC-RISK-SCREEN, these survival functions are fitted to 5 year survival for tumours of different stages ^30^.

### Model inputs

The same types of model inputs were specified in MANC-RISK-SCREEN as those used in the Gray-model. In some cases, however, new evidence was used to assign values to the model inputs. Tables 2, 3 and 4 provide an overview of key model inputs. Further details of how input parameters were calculated can be found in the model parameters and sources file in the documentation folder of the GitHub repository ^18^.

**Table 2:** Clinical parameters in base case model.

| Parameter | Value | Updated or added since Gray model? | Sources and Assumptions |
| --- | --- | --- | --- |
| Probability of attending a screening appointment | 60.5% for the first screen<br>85.2% if one previous screen attended<br>19.1% if first screen not attended | Yes | NHS Digital Screening and Immunisations Team, 2021<sup>40</sup> |
| Proportion of cancers detected by | 43.1% | Yes | Cancer Research |
| screening |  |  | UK, 2022 <sup>41</sup> |
| Age of death due to all-cause mortality | ~weibull(7.937,86.788) | Yes | Office for National Statistics <sup>42</sup> |
| Survival for stage 1 breast cancer | exp(-5.462) | Yes | Office for National Statistics 2019 <sup>30</sup> |
| Survival for stage 2 breast cancer | exp(-4.023) | Yes | Office for National Statistics 2019 <sup>30</sup> |
| Survival for stage 3 breast cancer | exp(-2.465) | Yes | Office for National Statistics 2019 <sup>30</sup> |
| Survival for stage 4 breast cancer for women aged up to 54 | exp(-1.787) | Yes | Office for National Statistics 2019 <sup>30</sup> |
| Survival for stage 4 breast cancer for women aged 55 to 74 | exp(-1.388) | Yes | Office for National Statistics 2019 <sup>30</sup> |
| Survival for stage 4 breast cancer for women aged over 75 | exp(-1.011) | Yes | Office for National Statistics 2019 <sup>30</sup> |
| Breast cancer incidence by age | See “Incidence_mortality_ONS2” input csv | Yes | Cancer Research UK 2022 <sup>31</sup> |
| 10 year risk of breast cancer estimated using the Tyrer-Cuzick version 8 and Volpara breast density group | See “synthetic_risk_data.csv” input | Yes | BC-PREDICT study |
| Lifetime risk of breast cancer estimated using the Tyrer-Cuzick version 8 and Volpara breast density group | See “synthetic_risk_data.csv” input | Yes | BC-PREDICT study |
| Volpara breast density estimate | See “synthetic_risk_data.csv” input | Yes | BC-PREDICT study |
| Probability that a cancer is metastatic given the age of the woman | 25=4.62%<br>35=8.67%<br>45=10.98%<br>55=12.71%<br>65=14.25%<br>75=15.98%<br>85=17.30% | No | Age nearest to woman’s current age at diagnosis of cancer used |
| Fraction of cancers that are ductal carcinoma in situ | 21.1% | Yes | NHS Digital Screening and Immunisations Team 2021 <sup>40</sup> |
| Probability of cancer being diagnosed as stage I, II, or III given size of tumour | See stage_by_size_mat in R script | Yes | Kollias et al. (1998), Wen et al. (2015), Cheng et al. (1997) <sup>36–38</sup> |
| Mammographic sensitivity by Volpara Density Group (VDG) | VDG1=85.0%<br>VDG2=77.6%<br>VDG3=69.5%<br>VDG4=61.0%<br>Average=75.7% | Yes | Wanders et al. (2017) <sup>34</sup> |
| Detection rate of mammography in high density screens | 4.2 per 1,000 screens | No | Tice et al. (2013) <sup>43</sup> |
| Recall rate for screening | 4.56% | Yes | Burnside et al. |
|  |  |  | (2018)<sup>35</sup> |
| Biopsy rate for screening | 2.40% | Yes | NHS Digital Screening and Immunisations Team (2021)<sup>40</sup> |
|  | Growth Model Parameters |  |  |
| Mean doubling rate for tumours | 4.12 | No | Weedon-Fekjær et al. (2008)<sup>27</sup> |
| Standard deviation of doubling rate for tumour | 3.93 | No | Weedon-Fekjær et al. (2008)<sup>27</sup> |
| Mean tumour doublings at clinical detection | 6.5 | No | Weedon-Fekjær et al. (2008)<sup>27</sup> |
| Standard deviation of tumour doublings at clinical detection | 0.535 | No | Weedon-Fekjær et al. (2008)<sup>27</sup> |
| Mean tumour doublings at screen detection | 6.12 | No | Weedon-Fekjær et al. (2008)<sup>27</sup> |
| Standard deviation of tumour doublings at screen detection | 0.96 | No | Weedon-Fekjær et al. (2008)<sup>27</sup> |
| Log normal mean of tumour growth rate | 1.07 | No | Weedon-Fekjær et al. (2008)<sup>27</sup> |
| Log normal standard deviation of tumour growth rate | 1.31 | No | Weedon-Fekjær et al. (2008)<sup>27</sup> |
| Maximum tumour size (mm) | 128 | No | Weedon-Fekjær et al. (2008)<sup>27</sup> |
| Starting tumour size (mm) | 0.25 | No | Weedon-Fekjær et al. (2008)<sup>27</sup> |

**Table 3:** Utility values used in the base case model.

| Parameter | Value | Updated or added since Gray model? | Sources and Assumptions |
| --- | --- | --- | --- |
| Age-related utility values for healthy individuals | 30=0.938<br>35=0.915<br>40=0.907<br>45=0.882<br>50=0.864<br>55=0.834<br>60=0.822<br>65=0.807<br>70=0.804<br>75=0.779<br>80=0.753<br>85=0.699<br>90=0.650 | No | Ara and Brazier (2011) <sup>45</sup> |
|  | 95=0.650<br>100=0.650 |  |  |
| Utility early breast cancer in the first year | 0.82 | Yes | Naik et al. (2017)<sup>48</sup> |
| Utility early breast cancer in subsequent years | 0.82 | Yes | Naik et al. (2017)<sup>48</sup> |
| Advanced breast cancer in the first year | 0.75 |  | Naik et al. (2017)<sup>48</sup> |
| Advanced breast cancer in subsequent years | 0.75 | Yes | Naik et al. (2017)<sup>48</sup> |

**Table 4:** Costs used in base case model.

| Parameter | Value | Updated or added since Gray model? | Sources and Assumptions |
| --- | --- | --- | --- |
| Cost of risk stratification | £8.69 | Yes | Wright et al (2023) <sup>12</sup> |
| Cost of mammography screen | £60.56 | Yes | Pragmatic microcosting<br>See model parameter update document |
| Cost of follow up | £106.16 | Yes | Inflated from value in Gray et al. (2017) <sup>1</sup> |
| Cost of biopsy | £290 | Yes | NHS England (2022) <sup>50</sup> |
| Cost of ultrasound screening | £52 | Yes | NHS England (2022) <sup>50</sup> |
| Cost of MRI screening | £114 | Yes | NHS England (2022) <sup>50</sup> |
| Cost of treating breast cancer by stage, age, and time lived with cancer | See tibble in R script | Yes | Laudicella et al. (2015) <sup>29</sup> |
| Cost of treating a ductal carcinoma in-situ | £9,480.15 | Yes | Inflated from value in Gray et al. (2017) <sup>1</sup> |

#### Cancer Incidence

Consistent with the Gray-model the probability that an individual would experience breast cancer in their lifetime was assumed to be equal to their predicted lifetime risk using the Tyrer-Cuzick questionnaire and Volpara automated breast density measurement. This assumption implies that the risk prediction strategy provides a perfect estimate of a woman’s risk of breast cancer. For women who experienced a breast cancer in the model, the age of incidence was applied based on the age distribution of cancers provided by Cancer Research UK ^31^.

#### Tumour growth model

The process for selecting tumour growth model (continuous time) is detailed in Gray et al and in the documentation folder of the GitHub repository ^1,18^. The chosen model was based on observations from Norwegian women, providing the closest approximation for women in the UK of the models investigated.

#### Breast Cancer Risk-Stratification

For each simulated woman in the model, three key risk parameters are recorded: Volpara breast density, 10-year breast cancer risk as defined by the Tyrer-Cuzick questionnaire including breast density, and lifetime breast cancer risk. To allow the sharing of this data in the model files, the synthpop package was used in R to generate a synthetic dataset which maintained the structure of the underlying data ^32^. For example, the process ensures that lifetime risk remains correlated to 10-year risk. In this analysis it is assumed that all women chose to have their breast cancer risk predicted and agreed to change their screening intervals. Parameters have been added to the model to allow for imperfect uptake of risk prediction in future analysis.

#### Uptake to screening

MANC-RISK-SCREEN builds on the Gray-model to allow imperfect uptake for breast cancer screening using three model parameters. Women have a 60.5% chance of attending their first mammography. For subsequent mammographies, the probability of attending is 85.2% if the woman has attended at least one previous screening event and 19.1% if she has never attended screening. All uptake data were taken from the NHS Digital annual breast cancer screening report ^33^.

The sensitivity of mammography is also dependent on the density of a woman’s breasts in that denser breast tissue can mask tumours, reducing the sensitivity of screening. Women are first assigned to a Volpara Breast Density group between 1 and 4 based on their breast density. To calculate the Volpara Breast Density group-specific sensitivity given the size of the tumour “the ratio of the odds of a true positive result for that Volpara Breast Density Group compared with the population average odds was combined with the odds of a true positive result given the tumor size alone” ^1^. The sensitivity of screening by Volpara Breast Density Group was updated using more recent evidence in MANC-RISK-SCREEN ^34^.

#### Mammography recall rate

Women who attend screening and do not have a cancer present can experience a recall for further testing in the model after receiving a false-positive result. The rate of recall was updated in MANC-RISK-SCREEN using data from the UK NBSP ^35^

#### Diagnosis and treatment by cancer stage

Upon diagnosis of a cancer in the model it is assigned to be an invasive, non or micro-invasive, or an advanced tumour according to the TNM staging system. This is based on the size of the tumour at diagnosis, taken from the tumour growth model. To assign cancers of a given size to different tumour types and stages, a matrix was created using data from Kollias et al 1999, Wen et al 2015 and Cheng et al 1997 ^36–38^. To estimate the TNM stage of a cancer, the degree of lymph node involvement must be ascertained in addition to the cancer size.

While Wen et al contains full details of the number of lymph nodes involved, Kollias et al only contains whether there was node involvement or not. As such the distribution of the number of nodes from Wen et al was applied to the data from Kollias et al to estimate the number of nodes involved for this data. When combined with data on non-invasive cancers from Cheng et al, this allows a cancer to be assigned to stage I, II, III, or non-invasive status with given probabilities based on its size. The probability of a cancer of a given size being advanced (metastatic) was taken from the NHS audit of screen-detected breast cancers ^39^.

### Health consequences

The relevant health consequences are life-years and quality-adjusted life-years (QALYs) gained, using the same approach as the Gray-model. To generate estimates of the life expectancy of a woman without breast cancer, updated national population life tables were used to estimate the parameters for a Weibull survival distribution ^44^. Life expectancy data were updated, using data taken from before the COVID-19 pandemic starting in February 2020. Survival estimates for breast cancer of stages I-III were taken from the five-year cancer survival rates estimated by the Office for National Statistics ^30^. Due to the limited availability of survival data, only exponential survival functions could be fitted to these estimates, although this was found to be the optimal choice of survival model in the early economic evaluation for survival by NPI grade ^1^. For stage IV cancers, exponential survival models were estimated in the same manner but for different age bands (38 to 54 years; 55 to 74 years; 75 to 99 years) to reflect the effects of age on cancer survival.

When a woman is diagnosed with cancer in the model, a new age of death is drawn using the survival function for her stage of cancer and age. In the event that this age of death is higher than the age of death without cancer, the age of death is re-drawn from a truncated distribution with a maximum at the all-cause mortality age of death. For screen-detected cancers, the new mortality age was calculated based on the time the cancer would have been detected clinically. This avoids a potential problem where women with similar-stage cancers detected earlier by screening also die at an earlier age.

The utility values to calculate QALYs were updated from those used in the Gray-model with the exception of the population norm utility values for different age bands that were sourced from Ara and Brazier 2011 ^45^.

Published utility values for different stages of breast cancer were sought from studies included in two recently published systematic reviews of breast cancer related utility values ^46,47^. From these reviews, two studies with value sets potentially relevant to the UK and model context were chosen.

The face-validity of the sourced utility values was assessed using a group of patient experts (three women) with experience of breast cancer. In an online meeting, facilitated by two researchers, the women were asked to choose which they thought best reflected the experience of women in the UK with breast cancer. The final chosen utility values were those from Naik et al (2017) comprising a utility value of 0.82 for cancer of stage I-III and 0.75 for stage IV cancers ^48^. The full process for choosing these utility values is detailed in the “parameter update version 0 to 1” documentation file of the model GitHub repository ^18^.

### Costs

As the perspective for this study is that of the UK relevant healthcare system, costs associated with screening, diagnosis and treatment are included in the model. All costs in the model were updated from those used in the original model. The cost of predicting a woman’s risk of breast cancer was estimated in a microcosting study using interviews with researchers and staff who had been involved in the BC-Predict trial ^12^(p3). This study estimated costs for three risk prediction strategies both in the trial setting and in the NHS. This evaluation uses the estimated cost for the Tyrer-Cuzick questionnaire and Volpara breast density measurement in the NHS (£6.64 per woman).

Accurate costs of mammography for breast cancer screening are not available in the UK, so a simple microcosting was conducted using input from the BC Predict study team. Full details of the calculation of mammography cost are available in the parameter update document included in the GitHub repository ^18^. The final estimated cost of mammography was £60.56. The cost of recall for false-positive results was inflated to 2021 values from those used in the Gray model. Updated costs of biopsy (£290) were taken from NHS reference costs ^49^.

To identify treatment costs for different stages of breast cancer a systematic review as undertaken. Full details of this review can be found in the parameter update document in the documentation folder in the GitHub repository ^18^. Only one study which was relevant to the UK was identified ^29^. In this study, the authors estimated the cost of treating breast cancer of either an early (I or II) or late (III or IV) stage for women under or over the age of 65.

Treatment costs were also identified for the year of diagnosis and up to 9 years post-diagnosis. These data were used to create an exponential regression predicting the total discounted treatment cost for a woman given her age of diagnosis, cancer stage at diagnosis, and life expectancy. As the cost of treating non-invasive tumours was not included in Laudicella et al, the cost used in Gray et al was inflated to 2022 values ^1,29^.

### Data analysis

Consistent with the Gray-model, this analysis calculates the discounted (see Table 1 for rates) expected healthcare costs, life-years and QALYs for each strategy.

A fully incremental analysis was conducted to identify the most cost-effective approach. There is currently no specific threshold cost-effectiveness recommended by the UK National Screening Committee, so we assume a threshold of £20,000 per QALY gained in line with that used by NICE ^51^. In addition to costs and QALYs, the average number of screens attended by women in each strategy was also recorded to demonstrate the potential resource impact of different screening strategies. In an additional analysis reported in the supplementary materials, the base case analysis was also conducted using a discount rate of 1.5% for both health outcomes and costs.

To explore the impact of parameter uncertainty on the optimal choice of screening, probabilistic analysis was undertaken. The distributions chosen for the parameters are shown in the supplementary materials table S1.1. Due to the number of simulations required to produce stable estimates, it was not feasible to run a large number of Monte-Carlo simulations for the probabilistic analysis. Instead, Monte Carlo simulations were conducted using wide distributions for the parameter estimates and generalised additive models fitted to predict the costs and QALYs for each strategy given the values of the input parameters. A dataset of 1 million simulations of the parameter values was then generated using the realistic parameter distributions, and the GAM models used to estimate costs and QALYs for each strategy for each parameter-set. The results of this analysis were then used to produce cost-effectiveness acceptability curves. Additional analysis, reported in the supplementary materials, was also conducted to determine the impact of changes in the parameter values on the optimal screening strategy.

## Results

The indiscriminate approaches with fixed intervals (including 3-yearly current NHS practice) are extendedly dominated, implying that we would always prefer risk-stratified strategies regardless of the value we place on QALYs. Risk-2 (basing screening frequency on tertiles of predicted 10-year risk) is the most effective and cost-effective approach; that is, it produces the most QALYs, and does so at a marginal cost that narrowly meets a threshold of £20,000 per QALY. Risk-1 and Risk-3, which only differ by screening interval for low-risk women, have very similar results; however, the small amount of money saved by limiting screens is insufficient to offset the small loss in QALYs resulting from missed cases.

Although Risk-2 was the most cost-effective it was also very resource-intensive, requiring women to attend an average of 9.03 screens – more than double the number of screening events expected in current practice. Conversely, Risk-1 and Risk-3 only required women to attend an average of 4.90 and 4.67 screens respectively, an increase of only 0.42 and 0.19 screens compared to 3-yearly screening (4.48).

### Probabilistic Results

The results of the probabilistic analysis are shown in table 6. The cost-effectiveness acceptability curve in figure 2 illustrates the impact of assuming different thresholds for cost-effectiveness. At thresholds lower than £11,000 per QALY, no screening is the strategy most likely to be cost-effective. Risk-1 is then the most likely to be cost-effective at thresholds up to £25,000 per QALY, with Risk-2 becoming the most cost-effective at higher thresholds.

**Figure 1:**
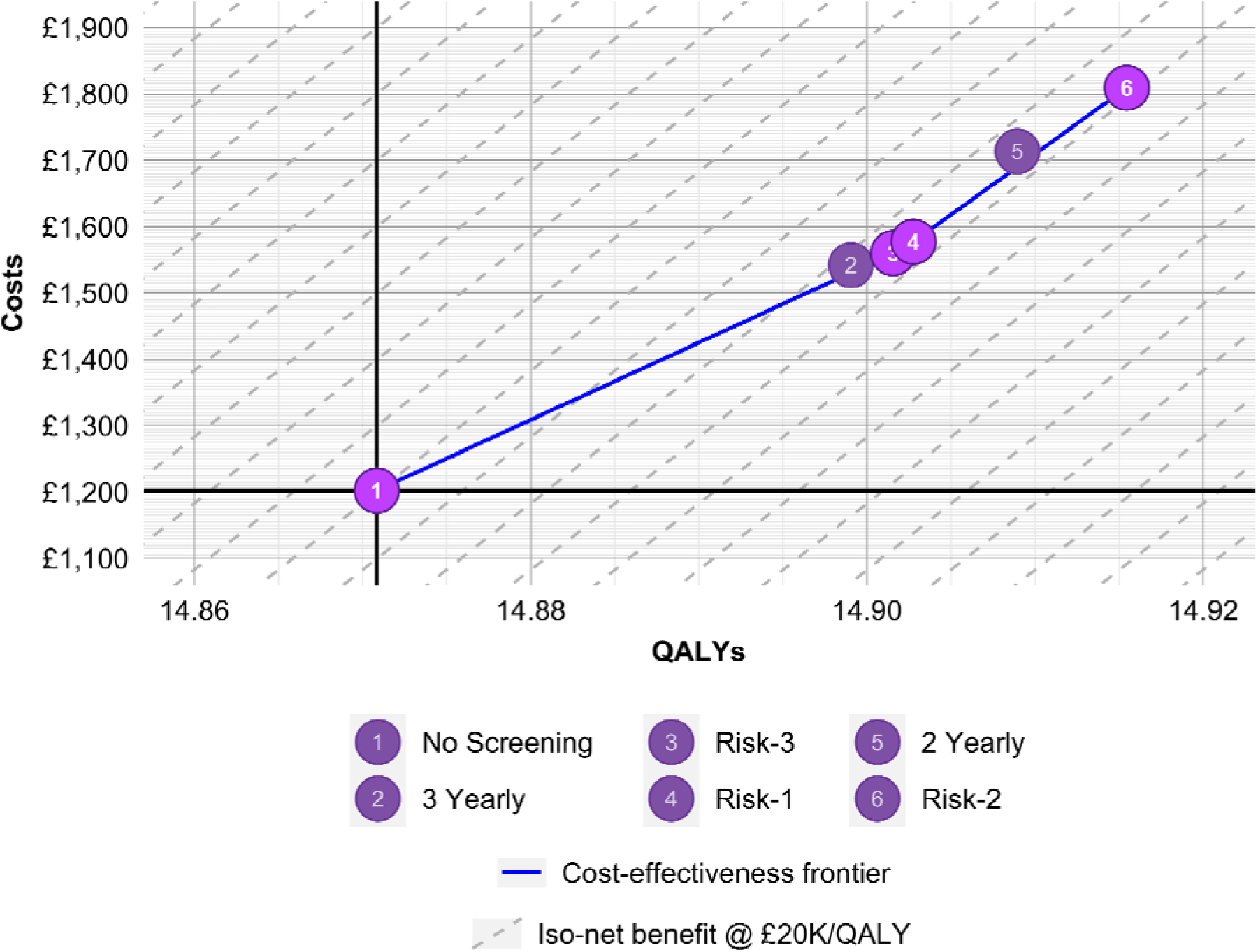
Cost-effectiveness frontier.

**Figure 2:**
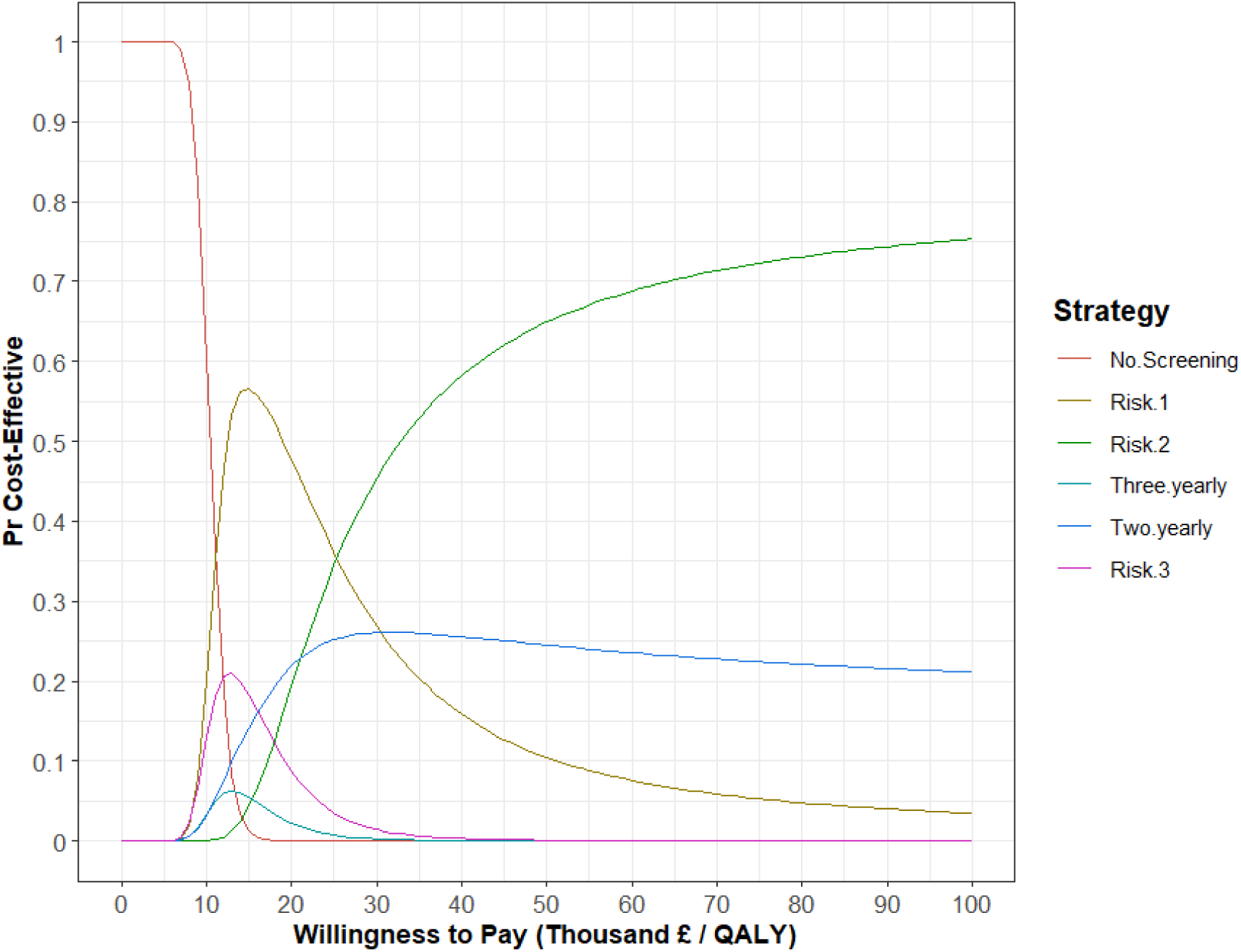
Cost-effectiveness acceptability curve.

**Table 5:** Base Case Results of MANC-RISK-SCREEN.

| Strategy | Costs (£)<sup>1</sup> | Effects (QALYs)<sup>1</sup> | Effects (Life Years)<sup>1</sup> | Screens | Incremental Costs (£) | Incremental Effects (QALYs) | Incremental Cost-Effectiveness Ratio | Net Health Benefit<sup>2</sup> | Rank |
| --- | --- | --- | --- | --- | --- | --- | --- | --- | --- |
| **No Screening** | £1202 | 14.871 | 19.672 | 0.00 | - | - | - | 14.8109 | 6 |
| **3 Yearly** | £1542 | 14.899 | 19.698 | 4.48 | £340 | 0.028 | Extendedly Dominated | 14.8219 | 5 |
| **Risk-3** | £1560 | 14.902 | 19.701 | 4.67 | £358 | 0.031 | £11643 per QALY | 14.8240 | 3 |
| **Risk-1** | £1577 | 14.903 | 19.702 | 4.90 | £17 | 0.001 | £14347 per QALY | 14.8242 | 2 |
| **2 Yearly** | £1714 | 14.909 | 19.707 | 7.44 | £136 | 0.006 | Extendedly Dominated | 14.8233 | 4 |
| **Risk-2** | £1809 | 14.915 | 19.714 | 9.03 | £232 | 0.012 | £19,333 per QALY | 14.8246 | 1 |

<sup>1</sup> All reported costs and effects discounted at a rate of 3.5%
<sup>2</sup>At a threshold of £20,000 per QALY

**Table 6:** Results of the Probabilistic Analysis.

| **Strategy** | **Costs (£)<sup>1</sup>** | **Effects (QALYs)<sup>1</sup>** | **Incremental Costs (£)** | **Incremental Effects (QALYs)** | **Incremental Cost-Effectiveness Ratio** | **Net Health Benefit<sup>2</sup>** | **Rank** |
| --- | --- | --- | --- | --- | --- | --- | --- |
| **No Screening** | 1200 | 14.865 | - | - | - | 14.805 | 6 |
| **3 Yearly** | 1543 | 14.892 | 343 | 0.027 | Extendedly Dominated | 14.815 | 5 |
| **Risk-3** | 1562 | 14.897 | 362 | 0.032 | Extendedly Dominated | 14.819 | 3 |
| **Risk-1** | 1577 | 14.900 | 378 | 0.034 | £10,964 per QALY | 14.821 | 1 |
| **2 Yearly** | 1700 | 14.904 | 122 | 0.005 | Extendedly Dominated | 14.819 | 2 |
| **Risk-2** | 1817 | 14.909 | 239 | 0.010 | £24,485 per QALY | 14.819 | 4 |

<sup>1</sup> All reported costs and effects discounted at a rate of 3.5%
<sup>2</sup>At a threshold of £20,000 per QALY

At higher cost-effectiveness thresholds the ordering of the probability of cost-effectiveness of different strategies aligns with the number of screens attended by the average woman. In clinical practice, resource-constraints in terms of the number of available radiographers or radiologists may make these strategies infeasible. Figure 3 presents a cost-effectiveness acceptability curve with Risk-2 (9.03 scans per woman) and 2-yearly (7.44 scans per woman) removed. In this case, Risk-1 is the most cost-effective strategy at thresholds over £11,000 per QALY.

**Figure 3:**
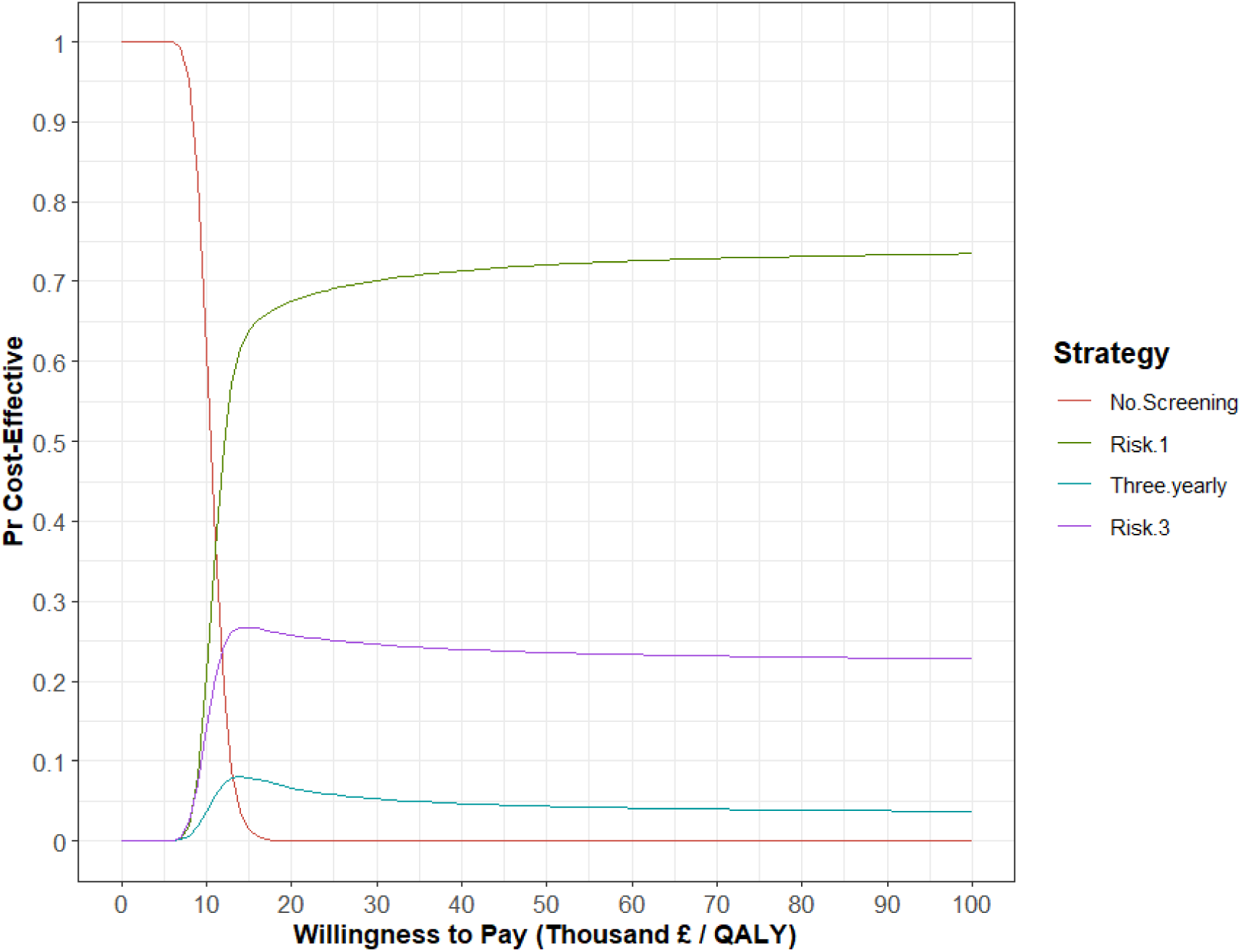
Cost-effectiveness threshold with resource intense strategies removed.

### Supplementary analysis

Additional analysis reported in supplementary materials S2 show the impact of changes in parameter values on the most cost-effective strategy at thresholds of £30,000 and £20,000 per QALY. More intensive screening strategies were more likely to be cost-effective when cancer survival, cancer utility and cancer growth rates were higher.

The results of the analysis when reducing the discount rate to 1.5% for both health outcomes and costs are shown in supplementary materials S3. In this scenario, the Risk-2 strategy became the most cost-effective at thresholds of both £20,000 and £30,000 per QALY. This was followed by 2-yearly screening and then the Risk-1 strategy,

### Comparing the results of the Gray and MANC-RISK-SCREEN models

Due to changes in the strategies evaluated and adaptations to the model structure, it is not possible to produce a quantitative comparison of the outputs of MANC-RISK-SCREEN compared with the original Gray model. Some general observations can be made. The expected costs of the different strategies were significantly higher in MANC-RISK-SCREEN. In contrast, the estimated expected QALYs were lower. These effects were likely to be driven by higher updated treatment costs and the change to only counting and discounting the costs and outcomes from the age of 50 rather than 38 years.

## Discussion

This study has reported an update of an early model-based cost-effectiveness analysis of risk-stratified breast cancer screening that we now label as a complete economic analysis ^16^. The original Gray-model and MANC-RISK-SCREEN produced similar results despite the latter using a different structure with updated parameter values. These results suggest that risk-based screening strategies are likely to be more cost-effective than universal breast cancer screening strategies.

At cost-effectiveness thresholds above £11,000 per QALY, there is always a risk-stratified breast cancer screening that is better than universal screening regardless of whether this is 2 or 3-yearly screening. In addition, screening involving more intensive screening was more cost-effective than less intensive screening at thresholds over £25,000 per QALY. Given the UK cost-effectiveness threshold of £20,000 per QALY and the existence of capacity constraints in the system such as the number of available radiographers, radiologists, and therefore mammograms, a risk-stratified approach which enhances cancer detection while minimising the number of additional screens is likely to be an attractive strategy in the NHS. Although reducing screening for women at lower risk of cancer may help to reduce the number of screens, the inferiority of Risk-3 compared with Risk-1 suggests that care should be taken to ensure the reduction in health benefits outweighs the savings in resources. Given the potentially infinite number of combinations of risk prediction approach, risk thresholds, and screening intervals, further research is needed to identify the strategy which maximises net benefit for a constrained number of screens.

The results of this study support findings by Hill et al that risk-based screening approaches are likely to be superior to 3 yearly screening in a UK NBSP ^13^. However, it is difficult to directly compare the results of the model as while the Risk-1 and Risk-2 strategies are similar to the ASSURE 1 and ASSURE 2 strategies in Hill et al, the MANC-RISK-SCREEN model uses updated risk thresholds for the moderate risk group (5-8%) while the Hill model uses the thresholds used in the Gray model (3.5-8%). For future evaluations of risk-stratified breast cancer screening in the UK, it would be useful to use the MANC-RISK-SCREEN, Hill et al, and Pashayan et al models to evaluate pre-specified stratified screening strategies in order to directly compare the results of the models.

### Limitations

The validation of the model revealed some weaknesses in its ability to produce outputs that match those observed in reality ^14^. While the distribution of stage of tumours identified at screening was similar in the model predictions and observed values, across all cancers the model tended to generate too many stage III cancers and too few at stage I.

The model outlined in this study has demonstrated that using the Tyrer-Cuzick questionnaire and Volpara breast density estimates to predict risk may be a cost-effective use of resources which does not materially increase the number of mammography scans across the NHS. However, there are many approaches to risk-prediction that could be combined in a large number of ways to predict a woman’s risk. For example, the CanRisk approach has been developed by researchers at the University of Cambridge, building on the BOADICEA risk prediction model ^52,53^. Further research is needed to understand the most cost-effective approach to risk-prediction for risk-stratified breast cancer screening. This should involve a full incremental analysis such that the cost-effectiveness of different risk prediction approaches is compared against all other available approaches as opposed to just 3 yearly screening.

In previously published economic evaluations, the cost of mammography screening has been taken from NHS reference costs. However, in recent years the cost of mammography for screening has not been included in the reference costs and simply inflating the last available cost may provide inaccurate estimates. In this study, we used the results of a microcosting undertaken by speaking to researchers with experience of the screening programme.

Updated treatment costs for different stages of cancer are also required for future versions of the model. The data included in this analysis relate to resource use from 2001 to 2010. Since these dates, treatment pathways for breast cancer will have changed significantly, resulting in changes in the pattern of costs by cancer stage. The cost of many of these treatments, evaluated by NICE, is confidential, making it difficult to discern their true impact on total treatment costs. In addition, Laudicella et al only break down costs into early (stage I and II) and late (III and IV) cancer, limiting the granularity of data and only providing a significant benefit of screening when there is a stage shift from stage III to II. In addition, the costs of treating DCIS were not included in this paper and the values included in this study are even more out of date.

Due to the very small differences in estimated QALYs between the strategies, a large number of simulated women are required to produce accurate estimates in the analysis. Stability was achieved in the results for the base case analysis but there is less certainty in the stability of the results of the probabilistic analysis resulting in the use of the GAM model. This is particularly important when comparing the cost-effectiveness of Risk-1 and Risk-2 which only differ in the reduction of screening for women with a 10-year cancer risk of less than 1.5%. The authors are conducting further developmental work to improve the speed of the model, allowing for a greater number of model runs and greater accuracy in order to provide greater certainty in the evidence for reducing screening for those at low risk.

## Conclusion

This updated cost-effectiveness analysis suggests that risk-stratified breast cancer screening is likely to be cost-effective compared to universal screening strategies. Further research is needed to optimise the combination of risk-prediction approach, risk thresholds, and screening intervals.

## Data Availability

The model described in this paper and all required input data are publicly available via GitHub: https://github.com/stuwrighthealthecon/MANC-RISK-SCREEN/tree/v1.2 .

## Precis

Risk-based breast cancer screening strategies are likely to be cost-effective compared to universal screening in the United Kingdom National Health Service.

## Supplemental materials

One document containing parameter values for the probabilistic analysis and supplementary analysis restricting the results to feasible studies, using a 1.5% discount rate, and determining the impact of parameter variation on the optimal strategy.

## Authors’ contributions

All authors meet International Committee of Medical Journal Editors (ICMJE) criteria for authorship. SW formulated the research question, updated the model parameters, ran analyses and led the writing of the manuscript.

GR completed the checklists and contributed to writing the manuscript.

EG formulated the research question and contributed to writing the manuscript. AD contributed to updating the model parameters and writing the manuscript.

Lorna McWilliams contributed to the conceptualisation of the project and reviewed the manuscript David French contributed to the conceptualisation of the project and reviewed the manuscript

D Gareth Evans contributed to the conceptualisation of the project, provided data from the BC Predict study, and reviewed the manuscript

KP formulated the research question, provided advice on the design for the overall study, and produced a first draft of the manuscript. KP acts as guarantor for this work.

This manuscript has been read and approved by all the authors.

Role of the funder: The funder had no role in the design and conduct of the study; collection, management, analysis, and interpretation of the data; preparation, review, or approval of the manuscript; and decision to submit the manuscript for publication.

## Acknowledgements

We would like to acknowledge the input of Rob Hainsworth from The University of Manchester who contributed to the update and validation of the model. We would also like to thank Charlotte Kennedy and Graham Scotland from the University of Aberdeen who in using the model helped identify some minor errors in the code and areas for future improvement. We also want to acknowledge the expertise of Tom Jones who conducted an external technical verification of the decision-analytic model.

