## Supplementary Materials for "Evaluation of a Risk-Stratified National Breast Screening Programme in the United Kingdom: An updated cost-effectiveness analysis"

Supplementary Materials 1: Distributions used in the probablistic analysis

Table S1.1: Input parameters with sampling distributions and hyperparameters used in PSA

| **Parameter** | **Distribution** | **Hyperparameter 1** | **Hyperparameter 2** | **Updated or added since Gray model?** |
| --- | --- | --- | --- | --- |
| **Mammographic Sensitvity** | | **Mean** | **Standard deviation** |  |
| Beta 1 | Normal | 1.47 | 0.1 | No |
| Beta 2 | Normal | 6.51 | 0.5 | No |
| **VDG modifiers** | | $\alpha$ | $\beta$ |  |
| VDG 1 | Beta | 96 | 16 | Yes |
| VDG 2 | Beta | 298 | 86 | Yes |
| VDG 3 | Beta | 212 | 93 | Yes |
| VDG 4 | Beta | 61 | 39 | Yes |
| **Growth rate distribution** | | **Mean** | **Standard deviation** |  |
| $\alpha_{1}$ | Normal | 1.07 | 0.09 | No |
| $\alpha_{2}$ | Normal | 1.31 | 0.11 | No |
| **Survival post-BC** | | **Mean** | **Correlated draws** |  |
| $\gamma$ stage I | Multivariate normal (MVN) | -5.46 | See Table 5 | Yes |
| $\gamma$ stage II | MVN | -3.82 | See Table 5 | Yes |
| $\gamma$ stage III | MVN | -2.72 | See Table 5 | Yes |
| **Survival metastatic BC** |  | **Mean** | **Correlated draws** |  |
| $\gamma$ stage IV, age <55 | MVN | -1.79 | See Table 6 | Yes |
| $\gamma$ stage IV, age 55-74 | MVN | -1.39 | See Table 6 | Yes |
| $\gamma$ stage IV, age >74 | MVN | -1.01 | See Table 6 | Yes |
| **Utility weights** | | | |  |
| Early | 1-exp(MVN) | -1.71 | See Table 7 | Yes |
| Advanced cancer | 1-exp(MVN) | -1.39 | See Table 7 | Yes |
| **Costs** |  | **Mean** | **Standard Deviation** |  |
| Risk stratification | Log normal | 2.13 | 0.06 | Yes |
| Cost multiplier (independently drawn and applied for each other cost item) | Normal | 0 | 0.102 | Yes |

**Table S1.2: Covariance matrix for survival post-BC**

| 0.08878 |  |  |
| --- | --- | --- |
| 0.01819 | 0.00373 |  |
| 0.01866 | 0.00384 | 0.00395 |

**Table S1.3: Covariance matrix for metastatic survival**

| 0.01157 |  |  |
| --- | --- | --- |
| 0.00884 | 0.00705 |  |
| 0.00804 | 0.00613 | 0.00560 |

**Table S1.4: Covariance matrix for utility values**

| 0.00309 |  |
| --- | --- |
| 0.00446 | 0.00643 |

Supplementary materials 2: Cost-effectiveness of risk-stratified breast cancer screening with feasible numbers of scans.

Figure S2.1: Cost-effectiveness threshold with resource intense strategies removed

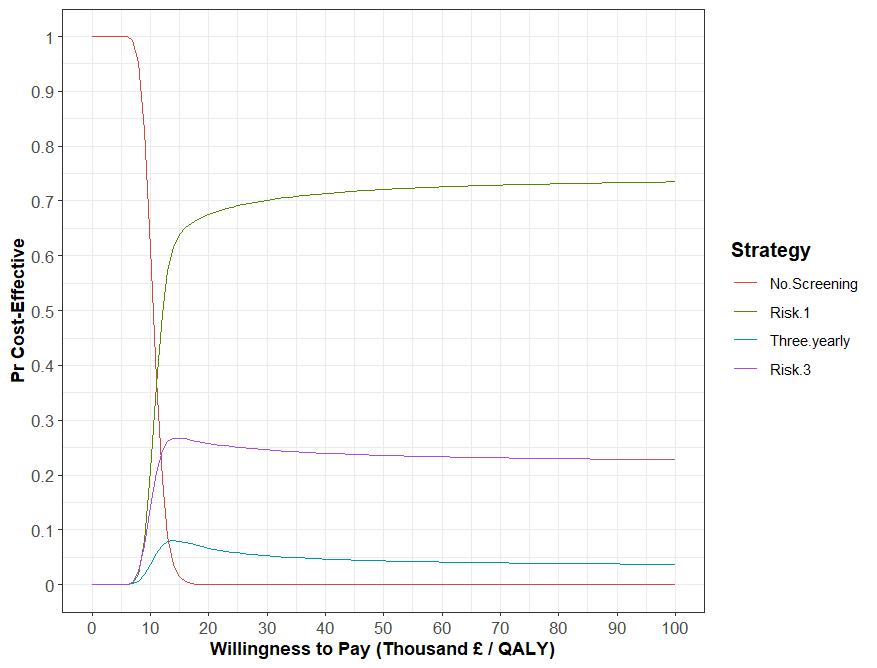

Supplementary materials 3: The impact of changes in parameter values on the most cost-effective strategy.

Figure S3.1: The impact of changes in parameter values on the most cost-effective strategy at a threshold of £30,000 per QALY

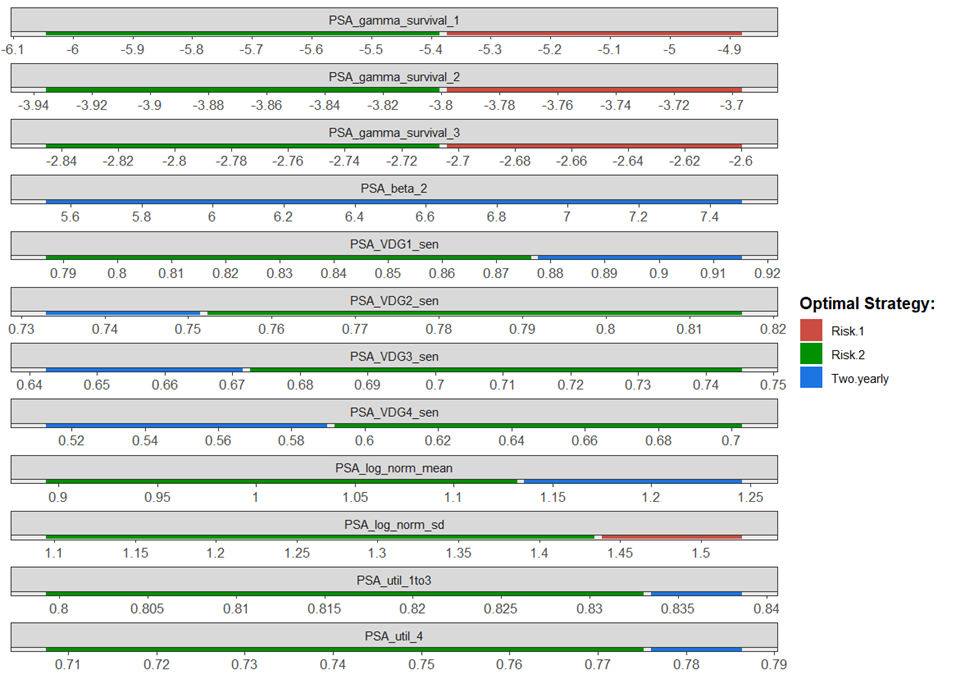

^1 PSA_gamma_survival_X=exponential survival curve parameter for stage X, PSA_beta_2=beta parameter for screening sensitivity logistic function, PSA_VDGX_sen=sensitivity of screening for volpara breast density group X, PSA_log_norm_mean=mean of log normal distribution for tumour growth rates, PSA_log_norm_sd=standard deviation of log normal distribution for tumour growth rates, PSA_util_1to3=utility values for tumours of stage I to III, PSA_util_4=utility value for tumour of stage IV^

At thresholds of £30,000 per QALY the risk-2 strategy was typically the most cost-effective except at low cancer survival rates where risk-1 was more cost-effective. Two-yearly screening was also more cost-effective than the risk-2 strategy when the sensitivity of screening was very high for VDG1 or very low for VDG2-4. 2-yearly screening was also the most-effective strategy when the mean tumour growth rate and utility values for stages I-III or IV was high. Changes in costs had no impact on the most cost-effective strategy.

Figure S3.2: The impact of changes in parameter values on the most cost-effective strategy at a threshold of £20,000 per QALY

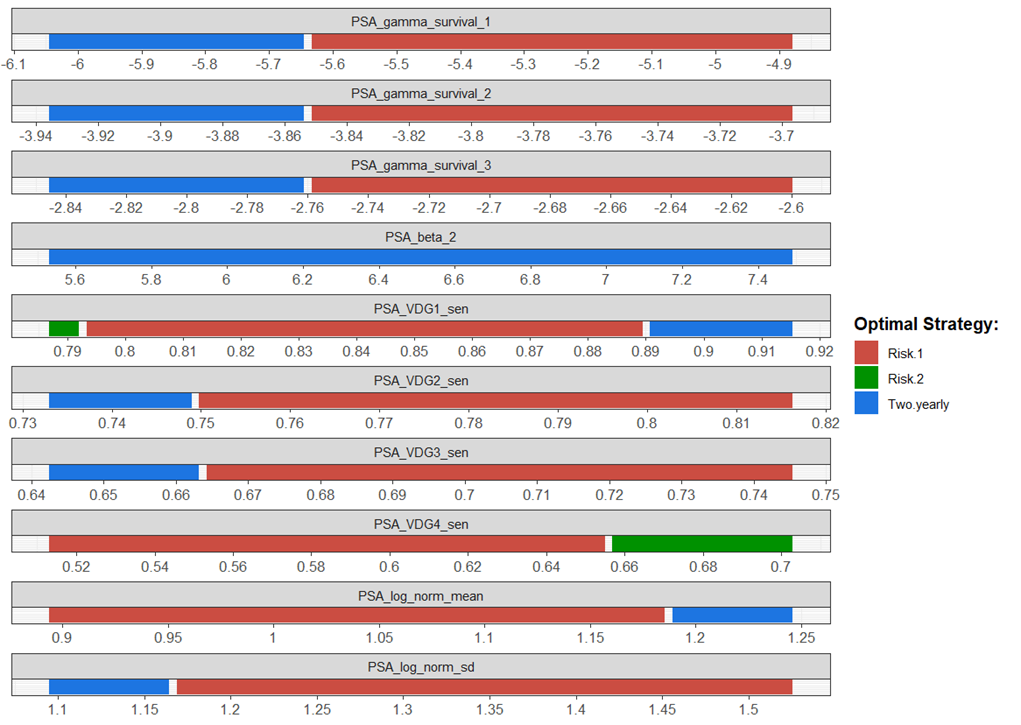

^1 PSA_gamma_survival_X=exponential survival curve parameter for stage X, PSA_beta_2=beta parameter for screening sensitivity logistic function, PSA_VDGX_sen=sensitivity of screening for volpara breast density group X, PSA_log_norm_mean=mean of log normal distribution for tumour growth rates, PSA_log_norm_sd=standard deviation of log normal distribution for tumour growth rates, PSA_util_1to3=utility values for tumours of stage I to III, PSA_util_4=utility value for tumour of stage IV^

At a threshold of £20,000 per QALY the risk-1 strategy was typically the most cost-effective. However, two-yearly screening was the most cost-effective strategy when survival rates were high or the mean tumour growth rate was high. The impact of variation in the sensitivity by VDG group was very mixed. Changes in utility values and cost had no impact on the results.

Supplementary materials 4: Results when using a discount rate of 1.5%

Table S4.1 shows the estimated costs, QALYs, and cost-effectiveness of the six strategies when the discount rate is reduced to 1.5% for both health outcomes and costs. Figure S3.1 shows the cost-effectiveness acceptability curve for this analysis.

Table S4.1: Costs, Outcomes, and Cost-effectiveness of Six Breast Cancer Screening Strategies

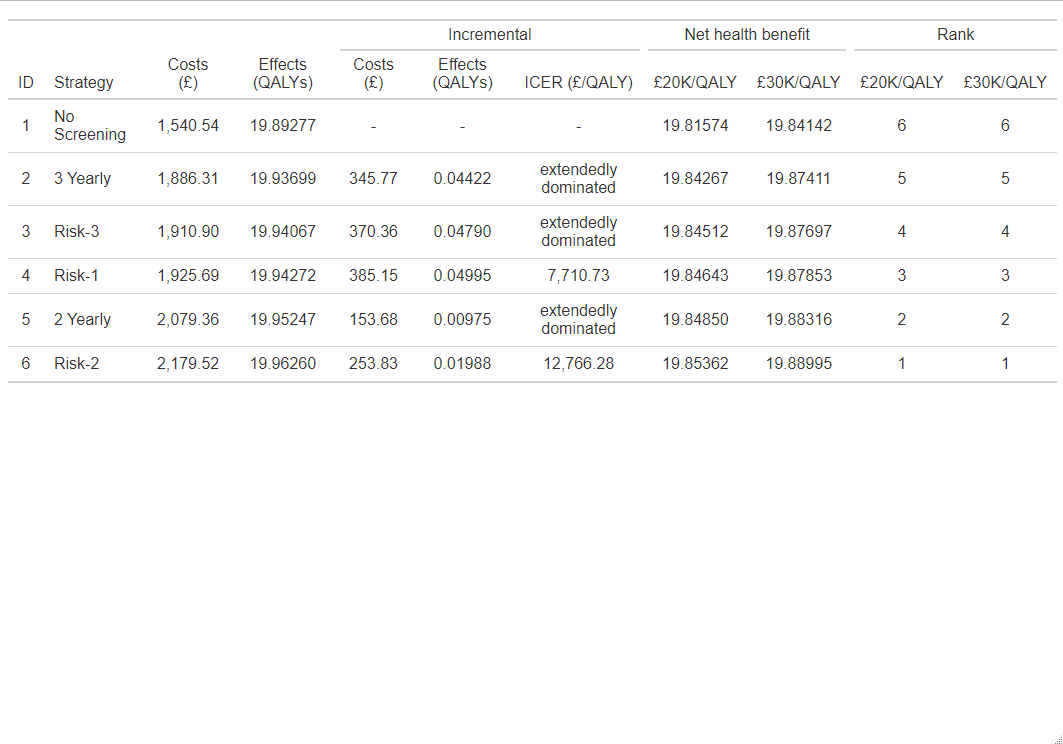

Figure S4.1: Cost-effectiveness Plane When Using a Discount Rate of 1.5%

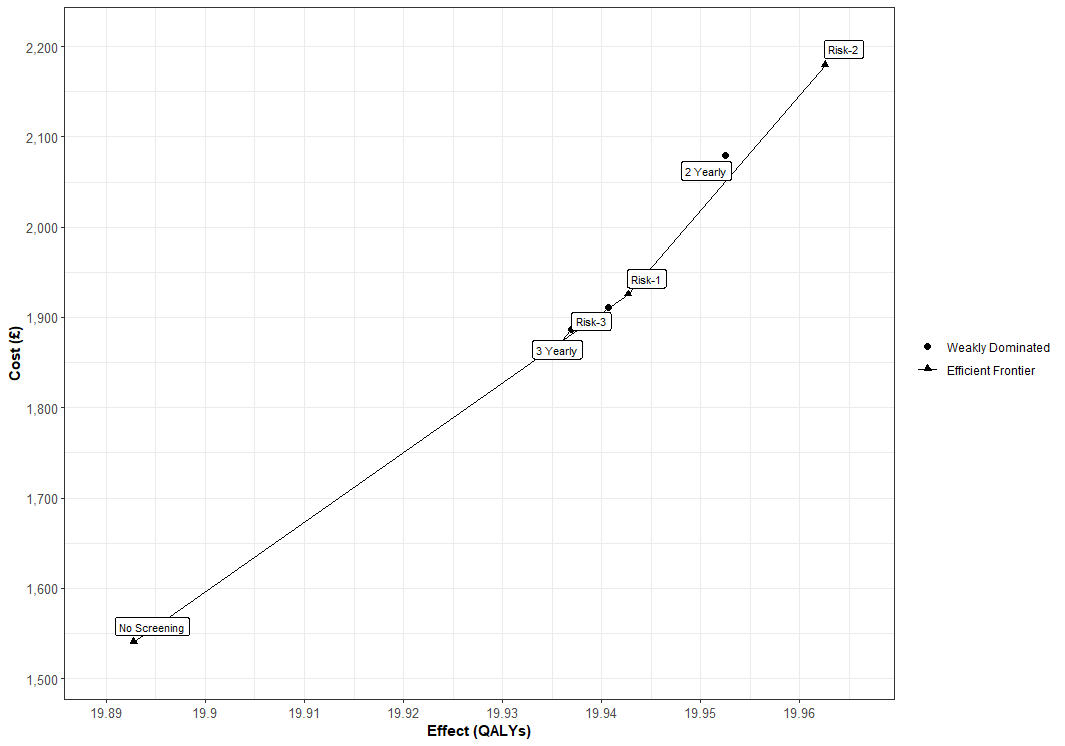
